# COVID-19 superspreading suggests mitigation by social network modulation

**DOI:** 10.1101/2020.09.15.20195008

**Authors:** Bjarke Frost Nielsen, Kim Sneppen

## Abstract

Although COVID-19 has caused severe suffering globally, the efficacy of non-pharmaceutical interventions has been greater than typical models have predicted. Meanwhile, evidence is mounting that the pandemic is characterized by superspreading. Capturing this phenomenon theoretically requires modeling at the scale of individuals. Using a mathematical model, we show that superspreading drastically enhances mitigations which reduce the overall personal contact number, and that social clustering (”social bubbles”) increases this effect.

---

During the ongoing COVID-19 pandemic, news stories have frequently appeared detailing spectacular events where single individuals – so-called *superspreaders* – have infected a large number of people within a short timeframe [1–3]. By now, there is substantial evidence that these are not just singular events, but that they reflect a marked transmission heterogeneity [4–6], a signature feature of the disease. In a well-mixed population, such heterogeneity has little bearing on the trajectory of an epidemic, but when public sphere contacts are restricted, heterogeneity takes on a decisive role, as shown in [7]. In that study it was demonstrated that superspreaders make the epidemic particularly sensitive to a reduction in random contacts as encountered in for example bars or large parties. In this report we investigate the effects of transmission heterogeneity – i.e. superspreading – on mitigation strategies which rely on a general reduction in social network size, and probe the influence of social clustering on such interventions.

The origins of superspreading can be diverse, depending on the characteristics of the pathogen in question. Superspreading events may occur due to circumstances and behaviour as well as biology. Even medical procedures, such as intubation and bronchoscopy, which facilitate the production of aerosols [11], can lead to superspreading events in respiratory diseases. However, the most straightforward model of superspreading is that some individuals simply shed the virus to a much greater extent than the average infected person. For COVID- 19, this *“biological superspreader”* phenomenon has some traction, and is supported by the observation that household transmission is limited, despite the relatively high *average* infectiousness of COVID-19 [12–14].

Superspreading is not a phenomenon which is particular to SARS-CoV-2, but has been observed in connection with several other pathogens, including coronaviruses such as SARS [15, 16] and MERS [9], as well as in diseases such as measles [8] and Ebola virus disease [17, 18]. Pandemic influenzas such as the 1918 Spanish flu, on the other hand, are believed to be far more “democratic” [10]. The heterogeneity of transmission is usually quantified using the Gamma distribution [8]. This is the origin of the *dispersion parameter* or *k value*, which determines the fraction of infectious individuals who account for the majority of infections (Fig. 1). Smaller *k* means greater heterogeneity – in fact, when *k* is small (|*k*| ≪ 1) it approximates the fraction of infected individuals who give rise to 80% of infections. For COVID-19, which is believed to have a *k* value of perhaps 0.1 [4–6], the most infectious 10% of individuals thus cause approximately 80% of infections.

**FIG. 1.**
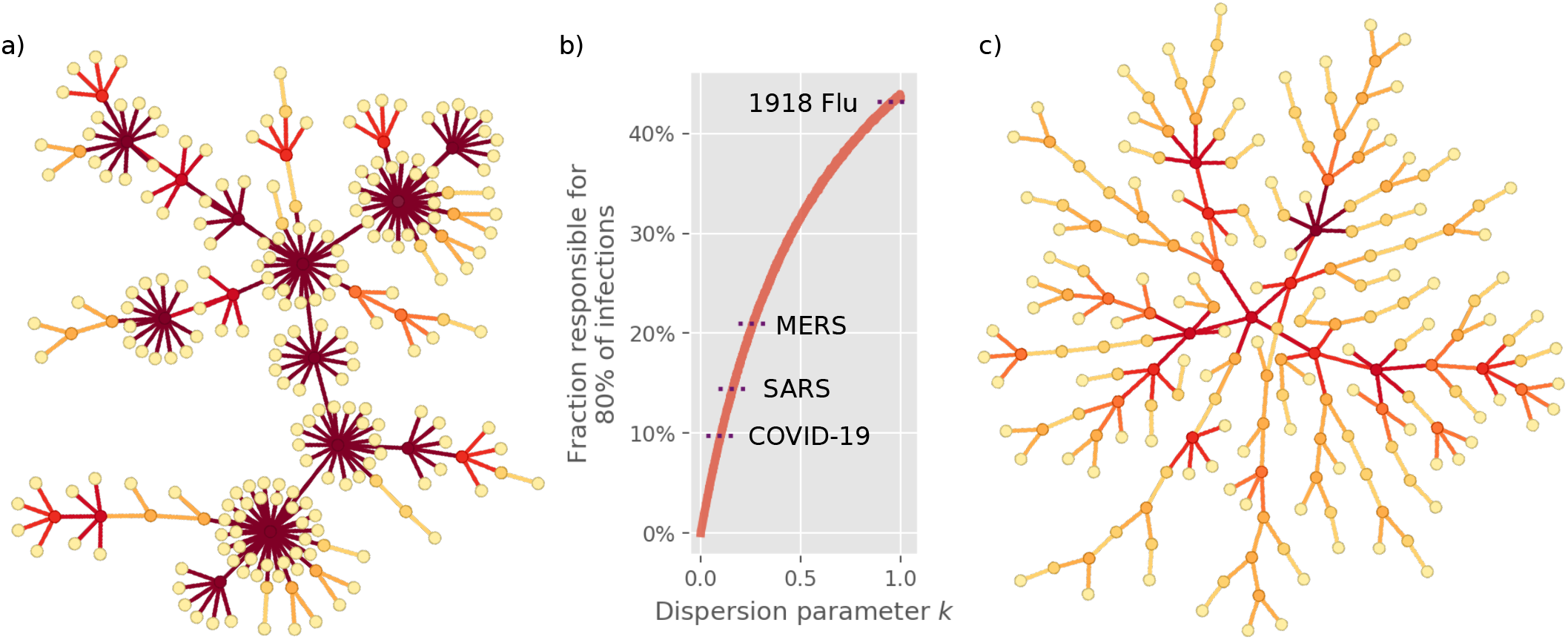
The characteristics of superspreading. **a)** Simulated infection network characterized by superspreading, with a dispersion parameter *k* = 0.1, within what has been observed for COVID-19 [4, 5]. Superspreaders appear as hubs, while most individuals are “dead ends” meaning that they do not transmit the disease. The epidemic mainly grows by spreading from one superspreader to the next. **b)** The dispersion parameter *k* provides a measure of superspreading, with lower *k* values corresponding to a greater heterogeneity. With a *k* value of 0.1 for COVID-19, approximately 10% of the population has the infectious potential to cause 80% of transmission. SARS and MERS are also characterized by a significant heterogeneity [8, 9], while pandemic influenza is believed to be more homogeneous [10]. **c)** Simulated infection network without superspreading (all individuals have equal infectious potential). Here, most individuals spread the disease to a few others, leading to a branched structure.

The fundamental difference between a homogeneously spreading disease and a highly heterogeneous one is reflected in the infection networks they give rise to, as visualized in Fig. 1. When only a small fraction of individuals cause the bulk of infections, a reduction in social network connectivity amounts to decreasing the likelihood that a superspreader infects another superspreader and thus propagates the disease. Consequently, in a network characterized by superspreading (Fig. 1a), the outbreak can be stopped by cutting only a few select edges. Not so for the network in Fig. 1c.

In this report, we present a model of superspreading phenomena which assumes that the driving force is a biological heterogeneity in infectiousness. We implement this as an agent-based model with contact networks, and are also able to capture much of the phenomenology in analytical formulae. In the model, *N* agents are placed as the nodes in a contact network. We investigate different types of network, but our base case is the Erdös-Renyi network, which is characterized by a Poisson degree distribution and an absence of clustering.

At initialization, the infectious potential of each individual is drawn from a Gamma distribution [8]. As such, it is an innate property of each individual. The possible states of each individual are **S**usceptible, **E**xposed, **I**nfected and **R**ecovered (see Suppl. Info. for details). At each timestep, each individual randomly selects one of its contacts to interact with, meaning that only a subset of the network is active at any given time. While a link between an infectious and a susceptible individual is active, there is a constant probability of infection per unit of time, as determined by the individual infectiousness.

This basic setup also lends itself to analytic calculations, as long as saturation effects can be ignored. Consider a single infected person who has *c* contacts. First, the infectious potential *r* of the individual is drawn from a gamma distribution *P*_*I*_ (*r*) with dispersion parameter *k* and mean *µ*. The distribution of the reproductive number *R* of an individual with a *known* infectiousness *r* and degree (i.e. connectivity) *c* is given by

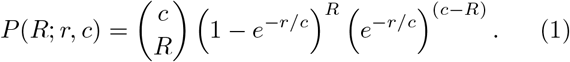

Taking the variability in infectiousness into account, the overall distribution of *R* becomes:

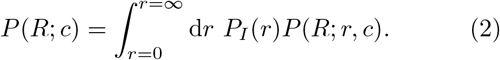

In the limit of infinite connectivity, corresponding to a well-mixed population, this becomes a negative binomial distribution. That particular case has been studied in [8]. Given a contact network, and a corresponding degree distribution *P*_*C*_(*c*) – for example a Poisson distribution in the case of an Erdös-Renyi network – the connectivities can be summed to yield a distribution of individual reproductive numbers, *P* (*R*) = Σ_*c*_ *P*_*C*_(*c*)*P* (*R*; *c*).

As reflected in the equations above, the *actual* number of secondary infections depends not only on biological infectiousness. In Fig. 2a-b, we explore how the number of personal contacts affects the resultant distribution of infections. Without superspreading (2a), a reduction in the contact number has a very modest effect and the distributions overlap. When the heterogenity is at a COVID-like level (2b), it is quite a different story. Here, a decrease in mean connectivity has a considerable effect, and mitigation suddenly looks feasible.

**FIG. 2.**
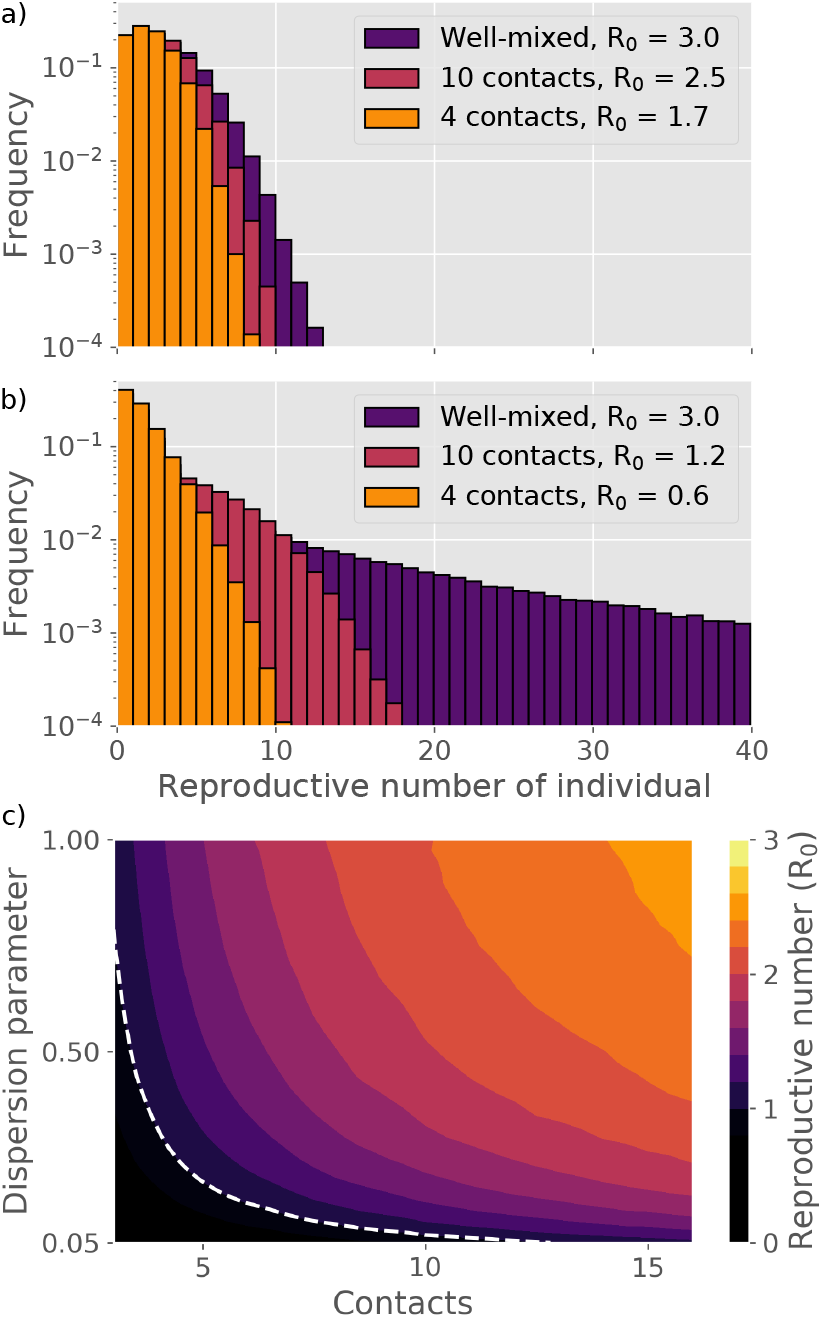
The reproductive number. Distributions of individual reproductive number *R* and value of *R*0 for different dispersion parameters and number of social contacts during an infectious period. **a)** Distribution of *R* for a disease where all individuals have equal infectious potential. **b)** Distribution of *R* for a disease characterized by superspreading (dispersion parameter *k* = 0.1). **c)** Basic reproductive number *R*0 as a function of social connectivity and dispersion. The dashed line represents *R*0 = 1.

To quantify the sensitivity of the epidemic to social network size, we consider the basic reproductive number *R*_0_, meaning the average number of infections that each infected person causes in a situation where all contacts are still susceptible. In Fig. 2c, the *R*_0_ is given as a function of the dispersion parameter *k* and the average contact number. The epidemic is evidently much more sensitive to reductions in average contact number when the transmission heterogeneity is high. A mitigation in which the average number of contacts goes from being unrestricted, down to about 10, causes a reduction in *R*_0_ which lowers both the *peak* and *total* number of persons infected during the course of the epidemic (the *attack rate*). The overall trajectory of a homogeneous disease is largely unaffected by social connectivity (Fig. 3a), whereas a heterogeneous epidemic is very sensitive (Fig. 3b). We find a particularly large sensitivity to a reduction of contact number from 15 down to 10 (Fig. 3b), indicating a critical threshold for disease spreading occuring at a contact number *∼* 1*/k*.

**FIG. 3.**
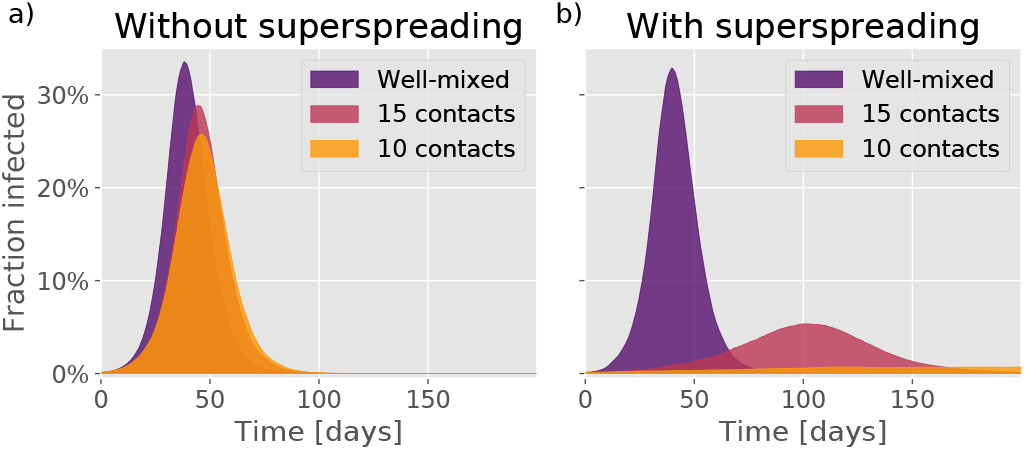
The epidemic trajectory of a heterogeneous disease is highly sensitive to mitigation. Epidemic trajectories as a function of the number of people that each person interacts with during an infectious period. **a)** Time evolution in the absence of any infection heterogeneity. **b)** Time evolution for a disease with dispersion parameter *k* = 0.1, roughly representative of COVID-19.

Crucially, a reduction in contact *time* is not necessary when the disease is characterized by superspreading. What counts is rather a reduction in contact *diversity*, meaning the number of different persons with whom you come into contact during the time you are infectious [7]. This differs fundamentally from SIR models, where contact time and diversity are not differentiated between [19, 20].

So far, our analysis has been based on the Erdös-Renyi network, which is largely devoid of clusters. This was chosen as a clean setting in which to probe how social network affects superspreaders. However, any realistic social network will involve clusters of people who know each other [21–23] – after all, your colleagues know each other, as well as knowing you. It is thus natural to ask whether such *cliquishness* impacts superspreading. In Fig. 4, we compare a cluster-free network to one characterized by a high degree of clustering [24].

**FIG. 4.**
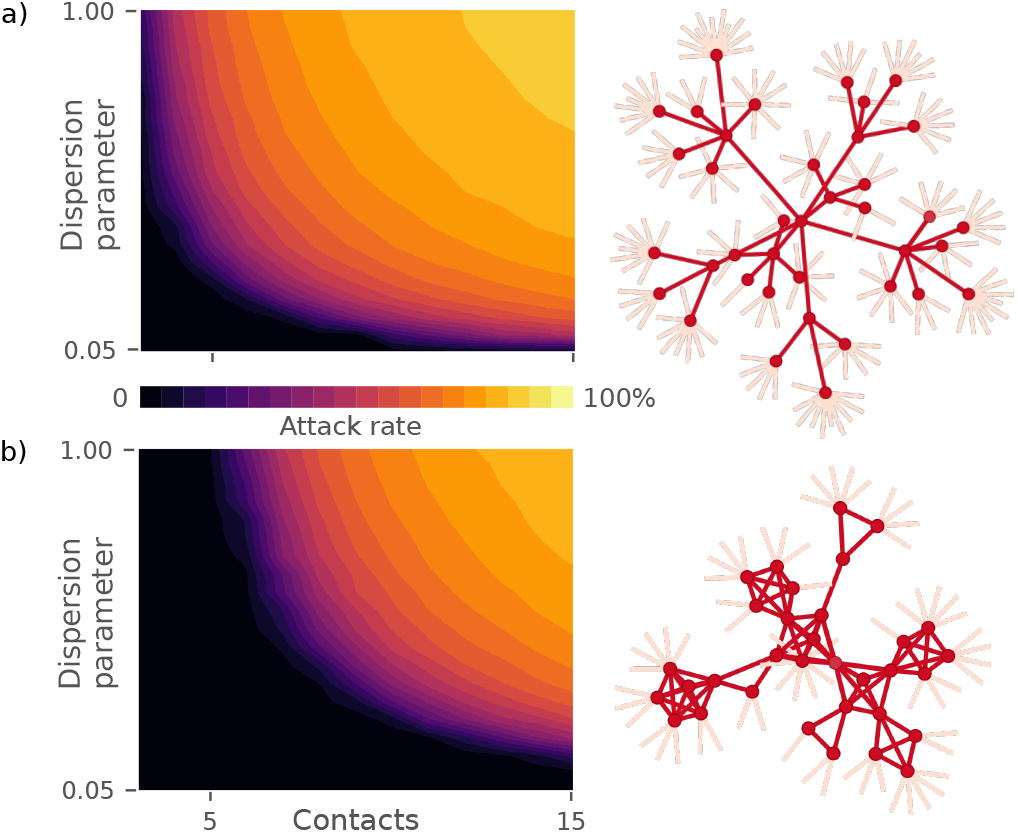
Final attack rate. (total fraction of the population infected) as a function of network connectivity and transmission heterogeneity. In panel **a)** we investigate the same Erdös-Renyi network as in Fig. 2. Panel **b)** explores a network where each person is assigned to two groups of people, leading to a highly clustered network. The black regions indicate conditions where the disease cannot spread in the population. On the right-hand side, small fragments of the networks in question are shown.

The attack rate of the disease is clearly lowered by clustering in general (Fig. 4), but the effect is especially significant when heterogeneity is high. The mechanism behind this is that of *local saturation*. If a superspreader infects a significant portion of his network, there is a risk that one of these individuals will turn out to be another superspreader. However, *if* there is clustering, a large part of this *second* superspreader’s network will already have been exposed, and the second superspreader does comparatively little harm.

Superspreading is now a well-established phenomenon for a number of diseases [8], including COVID-19 [4, 5]. In spite of this, the extent to which circumstance and person-specific properties contribute to the observed overdispersion in COVID-19 is still not clear. Super-spreading can also have a social component, exemplified by highly social individuals, who come into contact with a large number of people in a limited time-frame. However, such individuals would also be super*receivers*, a trait which impacts the epidemic even in the absence of mitigation [20, 25]. In any case, ability as well as opportunity is necessary for superspreading to occur. In our model, we have focused on inter-individual variation in ability to produce and transmit virus. This simplification is supported by cases of one person infecting many people at different times and locations [26], and by the observation that most infected people do not even infect their spouse [12–14]. However, more complex models could incorporate realistic *social* heterogeneity as well as large temporal variations in viral load [27] – effects which we have not probed. Furthermore, studies which address event-driven superspreading as well as contact tracing in the presence of superspreaders are also clearly needed.

Regardless of the origin of superspreading, we emphasize the particular fragility of a disease in which a major part of infections are caused by the minority. If this is the case, the disease is vulnerable to mitigation by reducing the number of *different* people that an individual meets within an infectious period. The significance is clear; Everybody can still be socially active, but generally only with relatively few – on the order of ten persons. Importantly, our study further demonstrates that repeated contact with *interconnected* groups (such as at a workplace or in friend groups) is comparatively less damaging than repeated contacts to independent people.

## Data Availability

The code developed for use in this study can be obtained from the authors by reasonable request.

## ACKNOWLEDGMENTS

We thank Lone Simonsen, Robert J. Taylor, Andreas Eilersen, Gorm G. Jensen and Julius B. Kirkegaard for enlightening discussions. Our research has received funding from the European Research Council (ERC) under the European Union’s Horizon 2020 research and innovation programme, grant agreement No. 740704.

