## Supplementary Information for "COVID-19 superspreading suggests mitigation by social network modulation"

### Supplementary Information: COVID-19 superspreading suggests mitigation by social network modulation

Bjarke Frost Nielsen* and Kim Sneppen†
*Niels Bohr Institute, University of Copenhagen.*
(Dated: October 4, 2020)

#### I. THE MODEL

The states of the model are shown in Figure 1, along with their durations (in days). Overall, the agents may be found in five states, **S**usceptible, **E**xposed, **I**nfected and **R**ecovered. The exposed state consists of two consecutive states with exponential waiting times (constant probability rate for leaving the state), thus forming a gamma distributed state when viewed as a whole. The infectious state is also divided into two states, shown as Presymptomatic and Infected. The two states are treated identically (save for their unequal durations), but are included for two reasons. First, it prevents unrealistically short infectious periods, which have a high probability of occurring if the state is modeled by a single constant-rate process, as in traditional S(E)IR models. Secondly, the presymptomatic state allows for straight-forward extension of the model to include e.g. contact tracing as well as differentiated infectiousness in the (pre-)symptomatic stages.
We have made a conscious choice to keep the model simple, to show the impact of superspreading directly, without introducing an excess of parameters.

The network simulation progresses in the following manner:

- Initialization:
  - A contact network is generated
  - For each agent, an infectivity is drawn from a distribution (Gamma distribution for superspreader scenarios, Dirac delta distribution for homogeneous scenarios).
  - The agents are initiated in the **S**usceptible state
  - A certain fraction (usually 0.1%) are designated as **I**nfected
- Time evolution:
  - For each node in the network, one attached edge is *activated*, simulating a contact event. This leads to highly connected nodes having multiple active edges in any given time-step.
  - If a network edge between an individual in an infectious state and a susceptible individual is activated, the susceptible individual has a probability rate for becoming exposed. The rate is given by the infectiousness of the infected individual.
  - Exposed individuals progress through the states at a rate given by the inverse of the average duration of the state.

In the case of a *well-mixed* simulation, a random contact is chosen, rather than choosing one from the agent's own contact network.

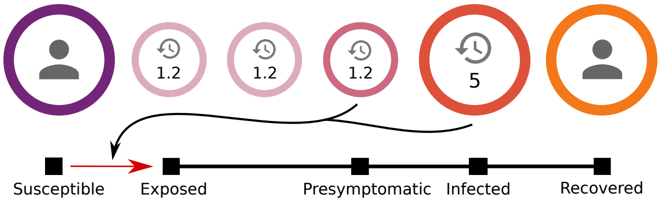


FIG. 1. The possible states of an individual in the model. The numbers indicate the average duration of each state, measured in days. The exposed state consists of two consecutive states with exponential waiting times, forming a gamma distributed state when taken as a whole. The infectious state is also broken into two states, shown as Presymptomatic and Infected.

*
†
